# Fear of Dementia among Middle-Aged and Older Adults in China: Lived Experiences, Coping, and Support Needs

**DOI:** 10.64898/2026.09.02.26361635

**Authors:** Huohuo Dai, Anke Versluis, Ruijie Xu, Wilco Achterberg, Hongxia Shen, Niels H. Chavannes, Jiska J. Aardoom

## Abstract

**Background:** Fear of dementia (FoD) can shape psychological well-being and responses to perceived cognitive change. Qualitative research has begun to show that dementia-related fear extends beyond memory loss to anticipated threats to independence, identity, and control. Less is known about how people manage these concerns in everyday life, what support they consider meaningful, and how these experiences unfold across sociocultural contexts. This study explored lived experiences of FoD, coping responses, and support needs among middle-aged and older adults in China.

**Methods:** In-depth semi-structured interviews were conducted on adults aged 45 years and older with some concern about developing dementia. A fictional third-person vignette facilitated discussion of this potentially sensitive topic. Data were analyzed using Framework Analysis, with Theoretical Domains Framework domains providing a deductive higher-level structure and more specific subthemes developed through iterative, data-driven comparison within and across cases.

**Results:** Twenty participants described fear of dementia as a socially embedded and future-oriented experience shaped by dementia-related knowledge, exposure, family roles, and concerns about autonomy, dignity, and burdening others. Coping strategies ranged from proactive self-management to avoidance and distraction, often supported by family members or peers. Participants consistently expressed a need for practical information, reassurance, and actionable strategies to maintain cognitive health. Preferred interventions were accessible, low-burden, and professionally guided. Although digital delivery was generally viewed as acceptable, engagement was perceived to depend on usability, trust, and digital literacy.

**Discussion and Implications:** Findings extend prior qualitative work on dementia-related anxiety by showing how fear is managed through both agency and avoidance and how support needs are embedded in relational and sociocultural contexts. Interventions addressing fear of dementia may benefit from being culturally sensitive, accessible, and responsive to users’ lived experiences. Attention to co-creation, participatory approaches, and equity may help support engagement and promote psychological well-being in ageing societies.

## Introduction

Dementia is a major global public health challenge, currently affecting more than 55 million people worldwide, with nearly ten million new cases diagnosed each year (World Health Organization, 2025). Population ageing is progressing rapidly, particularly in East Asian countries such as China. Meanwhile, concerns about memory loss have become increasingly salient across adulthood (Dai et al., 2023; Dai, Sun, et al., 2025), and a growing body of evidence demonstrates that dementia is one of the most feared chronic health conditions (Watson et al., 2023; Alzheimer’s Society, 2018).

Fear of dementia (FoD) refers to the emotional, cognitive, and behavioral responses associated with anticipating the possibility of developing dementia in the future, regardless of one’s current cognitive status (Kessler et al., 2012; Werner et al., 2021). Elevated FoD has been linked to negative consequences such as insomnia (Dai et al., 2023) and, at the severe end, anticipatory suicidal or death ideation (Maxfield, Peckham, James, et al., 2023). In addition to psychological distress, FoD may also relate to health-related avoidance behaviors, such as reluctance to undergo cognitive screening or seek timely medical care, which may hinder early detection and opportunities for intervention (Dai, Aardoom, et al., 2025; Farina et al., 2023). As societies confront an ageing population, understanding and addressing FoD may support psychological well-being, engagement in preventive health behaviors, and healthy ageing.

Although several interventions addressing FoD have emerged in recent years, the evidence base remains limited, and many interventions have been developed without a detailed understanding of how individuals experience, interpret, and manage dementia-related fears in everyday life (Dai, Aardoom, et al., 2025; Dai, Shen, et al., 2025). This matters because the relevance and acceptability of support may depend on whether it reflects the concerns people actually experience, the coping strategies they already use, and the social and practical contexts in which these concerns are managed. Understanding lived experiences is increasingly recognized as essential for developing interventions that are acceptable and relevant in real-world contexts (Kamarajah et al., 2026; The Lancet GlobalHealth, 2025). Qualitative research can therefore provide an important foundation for developing support that is responsive to lived experience rather than based primarily on assumptions about what people fear or need. (Kamarajah et al., 2026; Nevedal et al., 2025).

Existing qualitative research has begun to clarify why dementia is experienced as a particularly threatening prospect. Maxfield and colleagues found that community-dwelling adults without dementia described anxiety in relation to anticipated losses of independence, control, identity, and personhood, as well as future reliance on others (Maxfield, Peckham, & James, 2023). These findings suggest that FoD is embedded in how individuals imagine their future selves and relationships rather than being reducible to concern about memory loss alone. However, less is known about how people subsequently manage these fears in everyday life, why some respond through proactive self-management while others avoid or disengage, and what forms of support they consider meaningful and acceptable.

How these concerns are experienced and managed may also depend on the social and cultural contexts of ageing. In China, later-life independence is negotiated alongside strong intergenerational ties and expectations surrounding family care (Dai et al., 2023; Dai, Sun, et al., 2025). Concerns about future dependency may therefore be experienced not only as threats to personal autonomy but also in relation to reciprocity, family responsibility, and the possibility of burdening adult children. Stigma and norms surrounding emotional expression may further influence how dementia-related fears are discussed or concealed. Although quantitative studies have documented FoD among Chinese middle-aged and older adults (Dai et al., 2023; Dai, Sun, et al., 2025), little qualitative evidence has examined how these concerns are understood, managed, and translated into support needs in everyday life.

Addressing dementia-related concerns across diverse sociocultural contexts is increasingly recognized as a priority in global dementia research, which has been dominated by Western populations and high-income settings (Vilor-Tejedor et al., 2026). To address these gaps, this qualitative study aims to explore how middle-aged and older adults in China experience fear of dementia, the strategies they use to cope with these concerns, and their needs, preferences, and perceived barriers and facilitators regarding potential interventions. Specifically, the study seeks to answer the following research questions: (1) How do middle-aged and older adults experience and perceive FoD? (2) What strategies do they use to manage these concerns? and (3) What support needs and preferences do they describe, and what factors may facilitate or hinder engagement with potential interventions? By examining FoD across lived experience, coping, and support needs, this study seeks to extend existing qualitative work on dementia-related anxiety and inform culturally responsive, person-centered approaches to support and intervention development.

## Methods

### Study design

This study used an interpretative qualitative design to explore how middle-aged and older adults understood, experienced, and responded to FoD. A semi-structured interview topic guide was developed around the three research questions and used flexibly during data collection (***Appendix 1***). The Standards for Reporting Qualitative Research (SRQR) were used for reporting of this study (O’Brien et al., 2014) (***Appendix 2***).

### Study setting and participants

This study was conducted in urban and suburban communities across four provinces and one municipality in China between June 2025 and December 2025. The study settings were selected to capture a range of socioeconomic backgrounds in which concerns about cognitive ageing may arise.

Participants were middle-aged and older adults aged 45 years and above who reported concerns or fears related to developing dementia in the future. FoD was assessed using a single screening question: “How worried are you about developing Alzheimer’s disease in the future?”, rated on a 5- point scale ranging from 0 (“not worried at all”) to 4 (“extremely worried”). Individuals who scored 1 or higher were considered eligible for inclusion. A formal diagnosis of cognitive impairment or dementia was not required, as the study focused on anticipatory fear rather than clinical status.

Individuals with severe cognitive impairment or acute psychiatric conditions preventing meaningful participation were excluded.

Purposive sampling was used to seek variation in age, gender, education, living arrangements, and place of residence. Snowball sampling was subsequently used to identify additional eligible participants. Participants were recruited through community organizations, research assistants’ outreach, existing collaboration networks, and word-of-mouth referrals. Potential participants received a brief written or verbal explanation of the study, emphasizing that the research concerned experiences and worries related to memory and ageing rather than diagnostic assessment. This framing was used to reduce potential stigma and discomfort. Individuals who expressed interest were provided with detailed study information, given an opportunity to ask questions, and enrolled after providing written informed consent. Participants were offered RMB 30 as a token of appreciation for their time. The sample size was guided by considerations of information power (Malterud et al., 2016), including the focused study aim, specificity of the participant group, depth of the interviews, and planned cross-case analysis. Recruitment concluded when the research team judged that the dataset provided sufficient depth and variation to address the research questions.

### Data collection

Data were collected in settings familiar to the participants, including community centers, participants’ homes, or online platforms, according to participants’ preferences. Each participant completed a single, in-depth, semi-structured interview, lasting 35–55 minutes on average. Demographic information was collected at the start of the interview, and participants completed the Chinese version of the Fear and Avoidance of Memory Loss (FAM) scale (Dai, Sun, et al., 2025) for descriptive reference. The 17 item FAM comprises two constructs: fear and avoidance, with total scores ranging from 17 to 85, and higher scores indicating greater fear and avoidance of memory loss.

The research team included researchers with backgrounds in nursing, psychology, medicine, digital health, and qualitative health research across Chinese and European institutions. Interviews were conducted by four Chinese researchers with prior training and experience in qualitative research.

The interviewers had backgrounds in nursing and health research and included researchers at undergraduate-, master’s-, and doctoral-level training. Before data collection, all interviewers completed study-specific training to standardize their understanding of the study aims, research context, interview guide, use of the third-person vignette, interviewing procedures, and approaches to discussing potentially sensitive dementia-related concerns. Interviews followed a semi-structured topic guide covering participants’ experiences and perceptions of FoD, coping responses, and support needs and preferences (***Appendix 1***). Interviewers used neutral follow-up prompts where appropriate to clarify or deepen participants’ accounts.

Given the potentially sensitive and stigmatized nature of dementia-related fear, interviewers adopted a supportive and non-judgmental approach and reminded participants that they could pause or stop the interview at any time. A brief fictional third-person vignette describing “Auntie Li,” an individual experiencing dementia-related concern, was used as an initial conversational prompt. Participants were first invited to reflect on the vignette before being invited to relate the topic to their own views or experiences, if they wished. The vignette was intended to create psychological distance and reduce the immediate pressure of personal disclosure.

A separate observation form was used to document non-verbal expressions, interactional dynamics during and immediately after each interview, where applicable. All interviews were audio-recorded with participants’ permission, de-identified, and transcribed verbatim. Field notes recorded interviewers’ reflections on participants’ communication and engagement and were used to enhance the depth and contextualization of subsequent analysis where appropriate. None of the participants requested to review or amend their interview transcripts.

### Analysis

Descriptive statistics were used to summarize participants’ demographic characteristics and questionnaire data. Data were analyzed using Framework Analysis, following the Framework Method described by Gale et al., (2013). This approach was selected because the study addressed predefined areas of inquiry—experiences of FoD, coping, and support needs—while also requiring openness to unanticipated meanings and systematic comparison within and across participants. Analysis involved familiarization, coding, development and application of an analytical framework, charting into a framework matrix, and interpretation across and within cases (Gale et al., 2013).

We adopted a hybrid deductive–inductive analytical approach. The Theoretical Domains Framework (TDF) (Cane et al., 2012; Atkins et al., 2017; French et al., 2012) provided an a priori higher-level deductive structure for organizing cognitive, emotional, behavioral, social, and contextual influences relevant to how participants understood and responded to FoD. TDF domains were treated as analytical categories rather than predetermined findings. Within and across these domains, more specific codes and subthemes were developed inductively through iterative comparison of participants’ accounts. The analytical framework was therefore theory-informed while remaining open to patterns and meanings not adequately captured by the initial TDF structure.

Two researchers first familiarized themselves with the transcripts and recorded preliminary analytic observations and candidate codes. They jointly pilot-coded two transcripts to refine their shared interpretation and application of TDF domains and to identify inductive codes grounded in participants’ accounts. They then independently coded five additional transcripts and compared coding decisions. Differences were discussed to clarify code definitions, distinguish overlapping categories, and refine the preliminary analytical framework. Illustrative examples of how participants’ accounts were coded and incorporated into the analytical framework are provided in ***Appendix 3***. The refined framework was subsequently applied to the remaining transcripts. New inductive codes were added when participants’ accounts were not adequately represented by the existing framework, and previously coded transcripts were revisited when category definitions were refined. ATLAS.ti was used to support coding and data organization, while Microsoft Excel was used to construct framework matrices, with participants represented in rows and analytical categories in columns. The matrices facilitated systematic within-case and cross-case comparison, including attention to recurring patterns, divergent accounts, and relationships between participants’ interpretations of FoD and their coping responses.

The final findings were organized around the three research questions. TDF domains provided the higher-level deductive structure, while the reported subthemes reflected inductively developed patterns refined through iterative comparison across participants.

### Researcher reflexivity and rigor

Several strategies were used to enhance the credibility and transparency of the analysis. The two primary analysts maintained analytic notes documenting coding decisions, changes to the analytical framework, and emerging interpretations throughout the analysis. Independent coding and subsequent comparison were used to identify differences in interpretation rather than to seek statistical agreement. Differences were discussed to clarify code definitions, consider alternative explanations, and refine the developing analytical framework, with particular attention to accounts that did not fit dominant patterns.

Senior members of the research team reviewed the evolving analytical framework and interpretations during regular team discussions. These discussions were used to question assumptions, examine divergent cases, and consider whether interpretations remained sufficiently grounded in participants’ accounts. Framework matrices supported transparency by linking individual accounts to codes, TDF domains, subthemes, and cross-case interpretations. Reflexive discussions also considered how researchers’ disciplinary backgrounds and prior familiarity with dementia and FoD might shape interpretation. Reporting was guided by the SRQR (***Appendix 2***).

### Ethical considerations

The study was approved by the ethics committee of the relevant institution in China (reference number: 202504010). Participants received written or verbal information about the study’s purpose, procedures, potential risks, and benefits, and provided informed consent before participation. Participation was voluntary, and participants could withdraw or pause the interview at any time without consequence. Given the sensitive nature of discussing fear of dementia, interviewers adopted a supportive and non-judgmental approach. To compensate participants for their time, they were offered 30 RMB. Transcripts were de-identified to ensure confidentiality. Audio recordings and field notes were only accessible to the research team.

ChatGPT was used solely for language editing, including spelling and grammar checks, during manuscript preparation. All outputs were reviewed and revised by the authors, who take full responsibility for the final content. AI-assisted tools were not used for data collection, coding, analysis, or interpretation.

## Results

Participants’ socio-demographic characteristics are shown in ***Table 1***. Twenty participants were included, with a mean age of 59.0 years. Most were female (65%) and lived in urban areas (85%). Education levels varied from primary school (15%) to bachelor’s degree or above (35%). The majority were married (80%) and had two or more children (90%). Employment status and monthly income were diverse, and most participants had primarily resident basic medical insurance (70%).

**Table 1.**
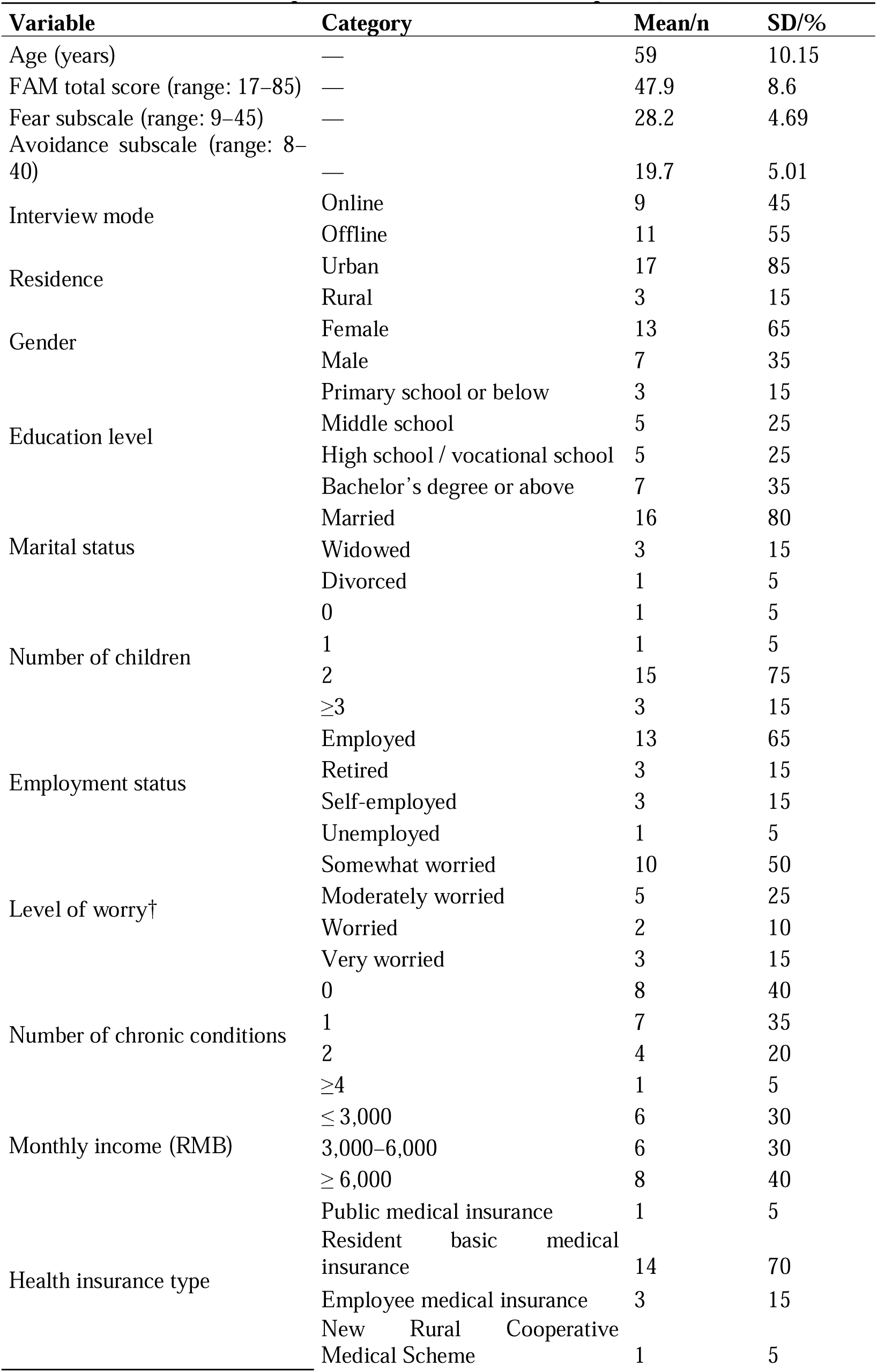
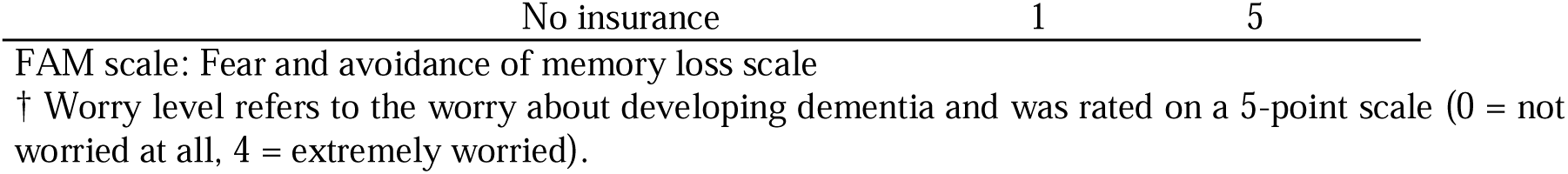
Descriptive Characteristics of Participants (N = 20)

The findings are presented according to the three research questions (***Tables 2–4***). Within each area, TDF domains are shown as higher-level analytical categories, with more specific subthemes developed through the iterative deductive–inductive analysis described above. Not all TDF domains were represented in the data; only domains supported by participants’ accounts are reported.

**Table 2.**
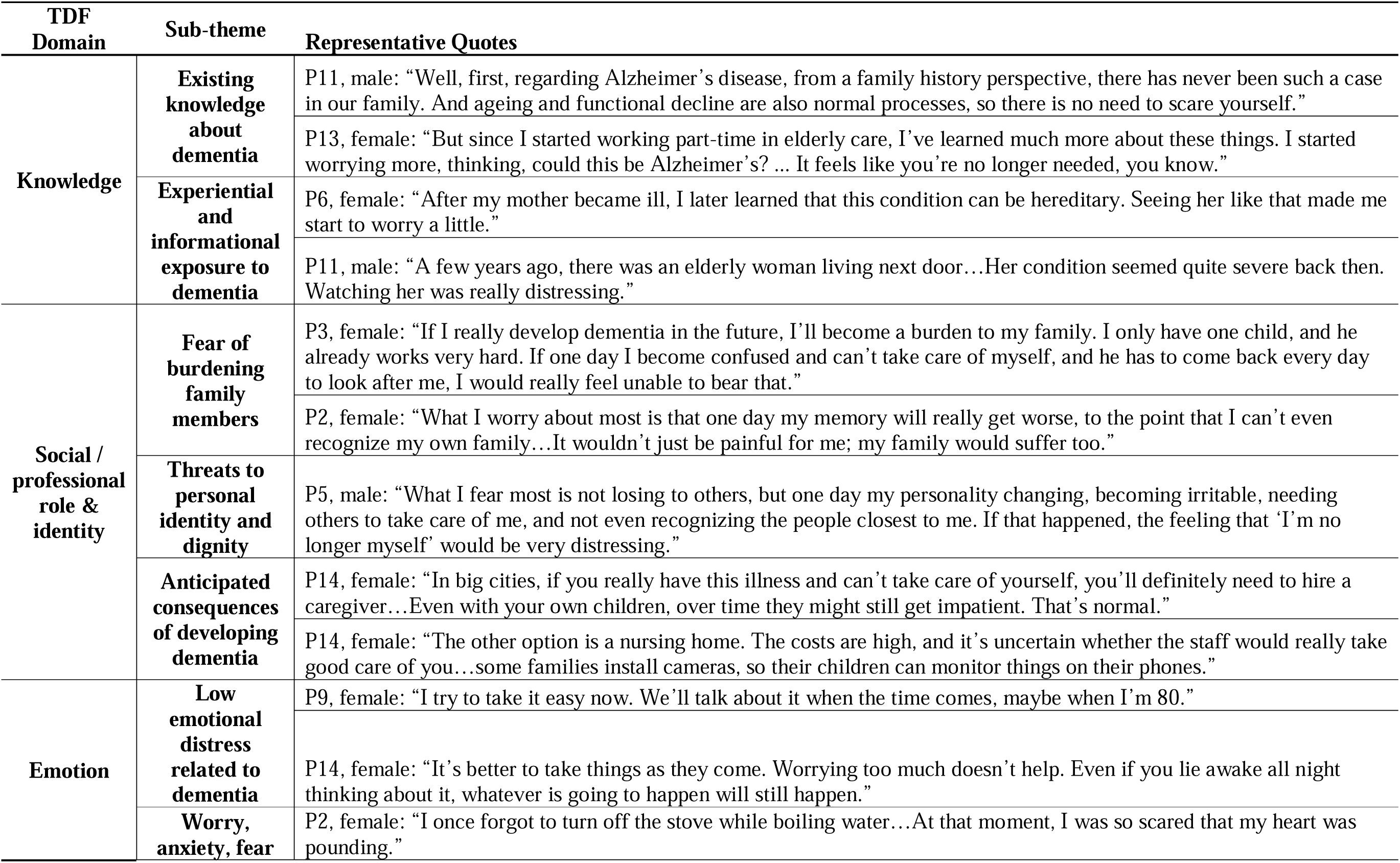
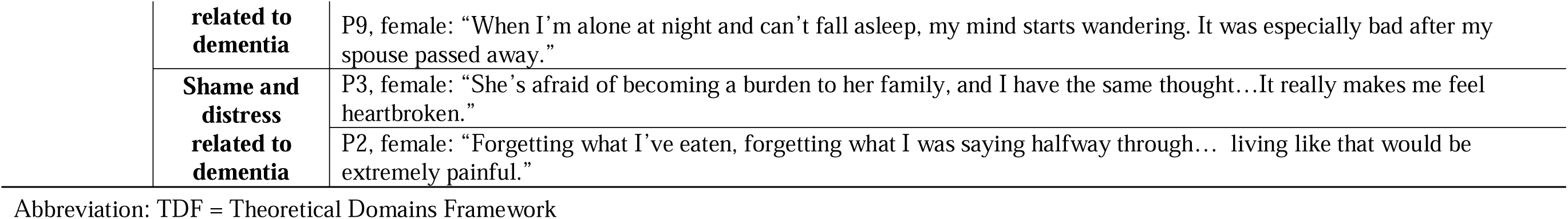
Experiences and Meanings of Fear of Dementia Representative Quotes.

**Table 3.**
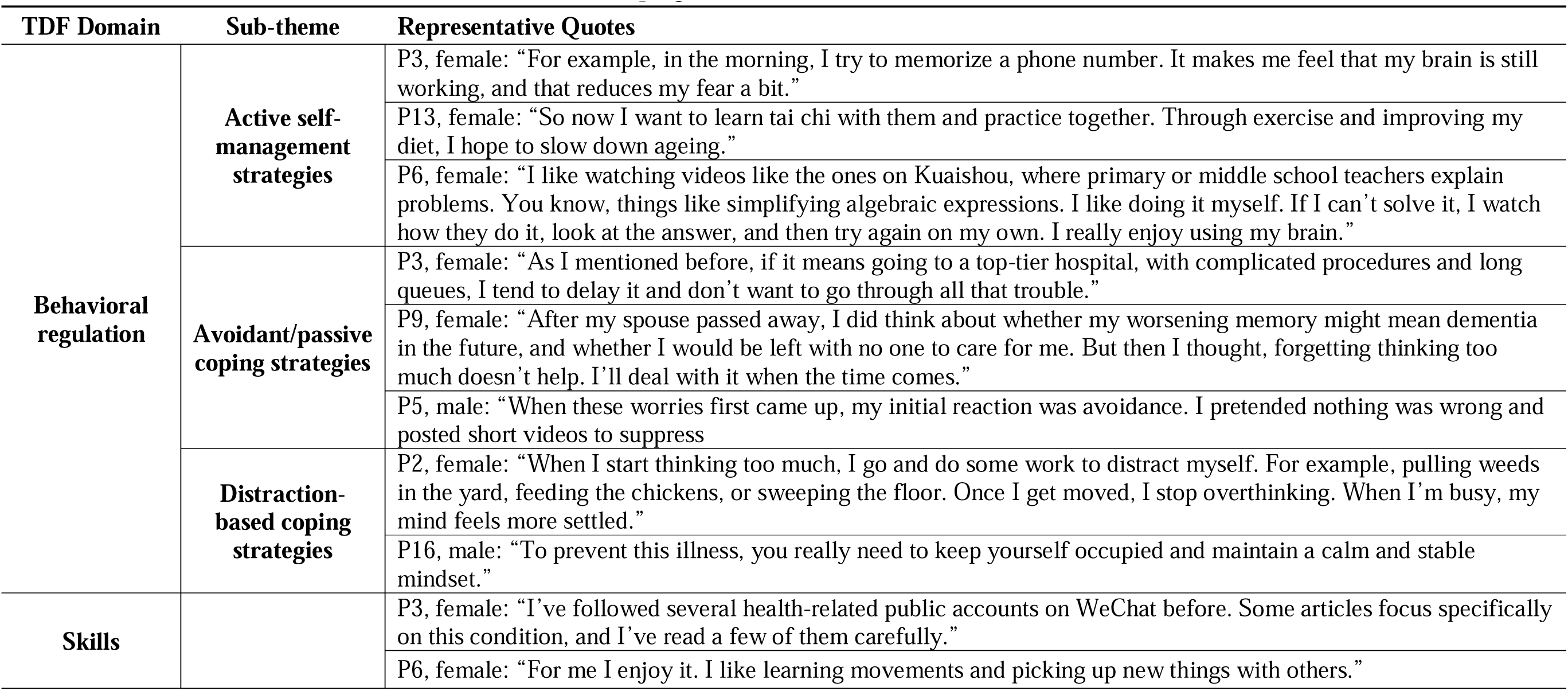
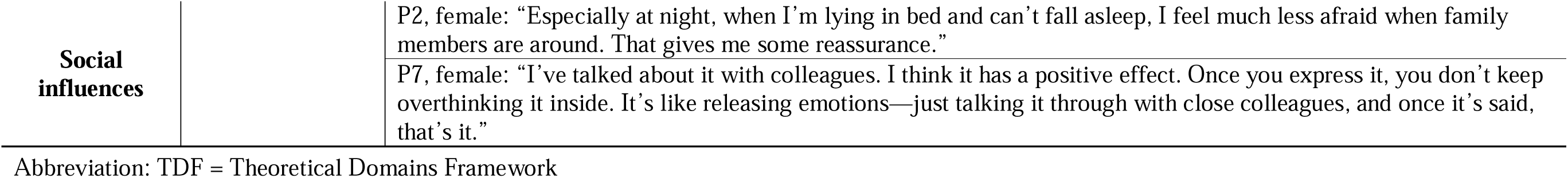
Coping With Dementia-Related Concerns TDF Domain Sub-theme Representative Quotes.

**Table 4.**
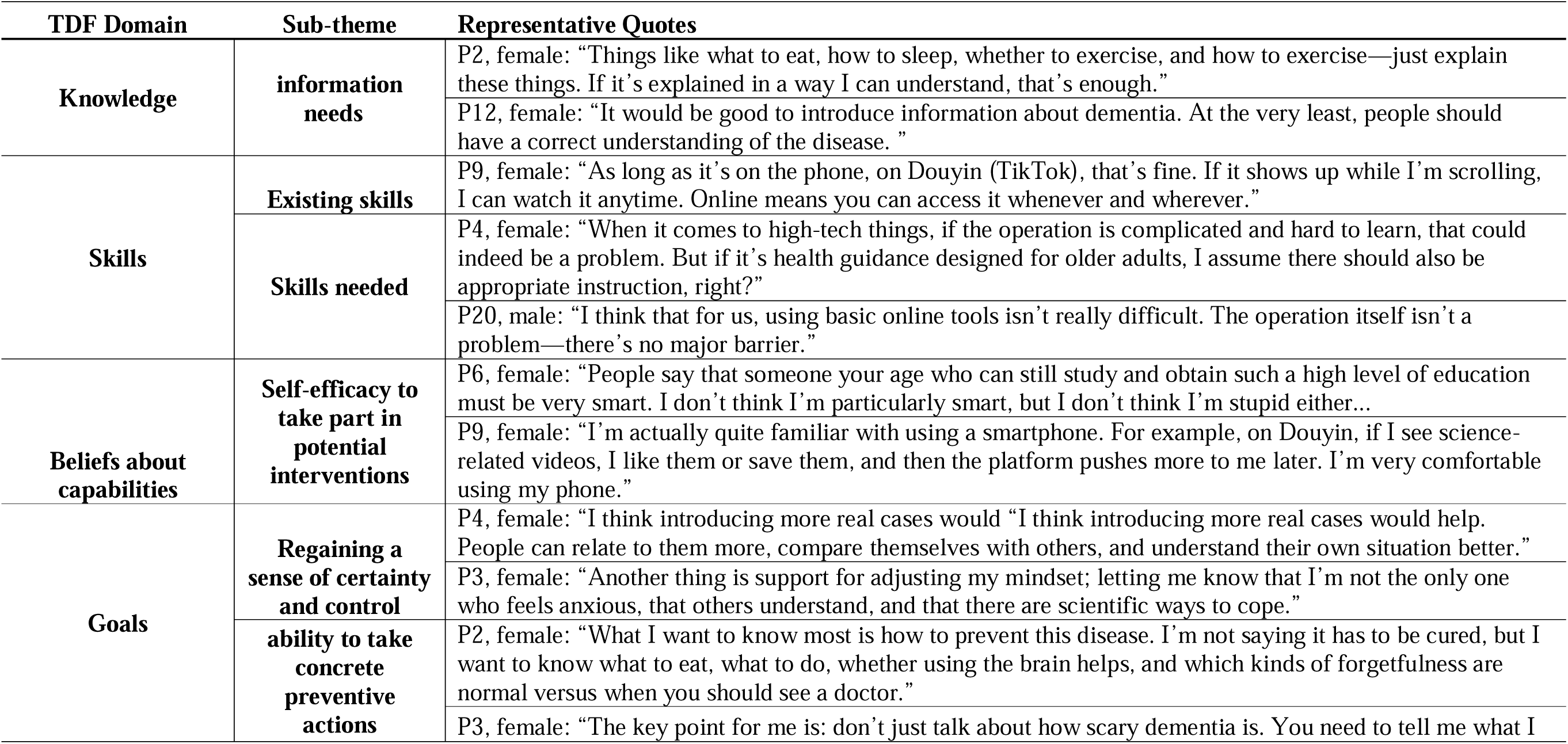
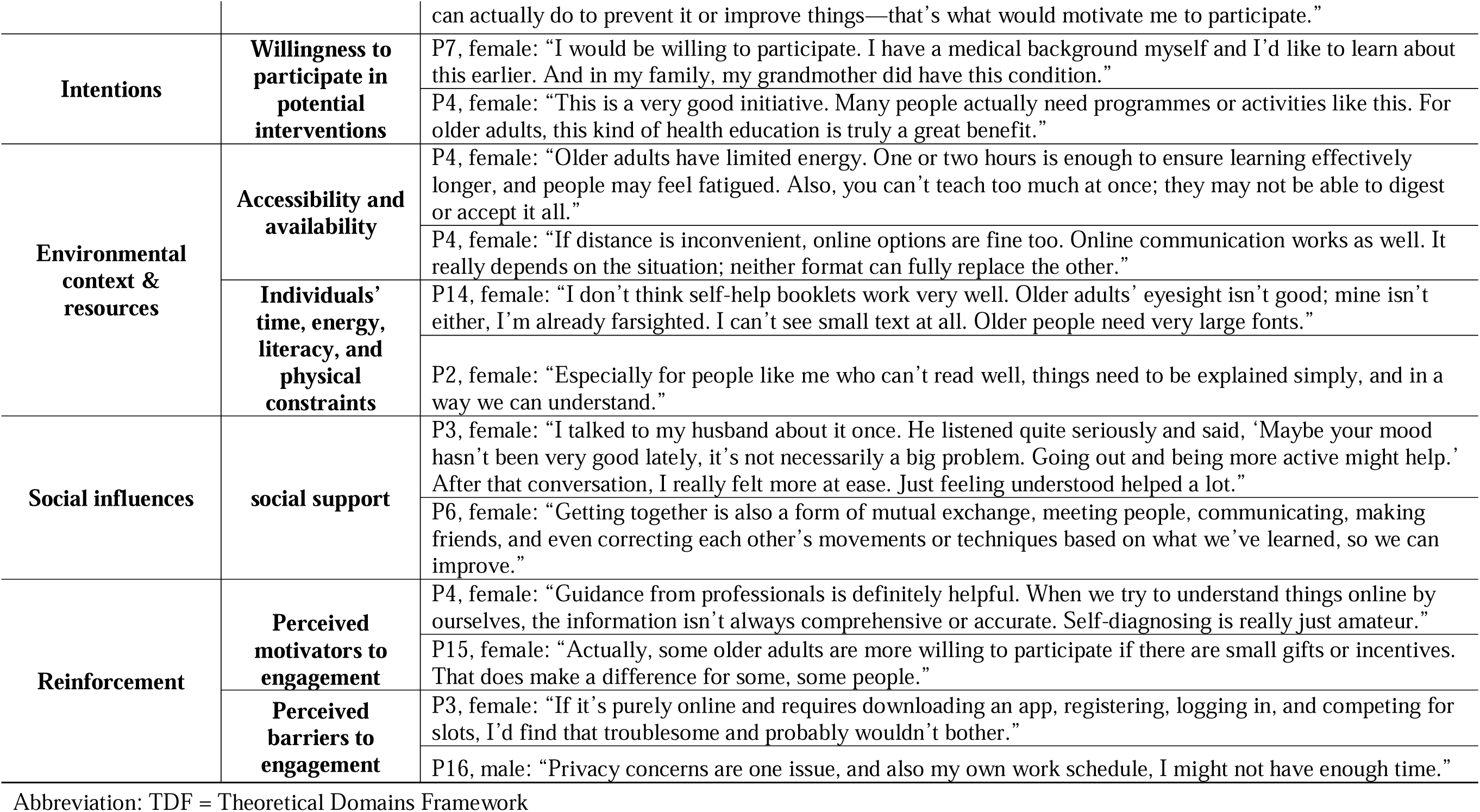
Support Needs and Preferences.

### Experiences and meanings of fear of dementia

Many participants drew on prior knowledge and experiences of dementia, particularly through family members, neighbors, or work contexts, which shaped their understanding of the condition and heightened their awareness of potential cognitive decline. While some perceived memory decline as a normal part of ageing and reported limited fear, others described how witnessing dementia in close others or learning about its hereditary nature intensified their worry and vigilance toward their own memory changes. For example, one participant reflected: *“After my mother became ill, I later learned that this condition can be hereditary. Seeing her like that made me start to worry a little” (P6, female)*.

These cognitive interpretations were closely intertwined with concerns about identity, autonomy, and social roles. A dominant theme was the fear of becoming a burden to family members, particularly adult children, often accompanied by worries about loss of independence and dignity. As one participant explained: *“If I really develop dementia in the future, I’ll become a burden to my family… If one day I become confused and can’t take care of myself, my child would have to come back every day to look after me” (P3, female)*. Beyond practical concerns, several participants expressed deeper fears about losing their sense of self or personhood, describing the possibility of personality change as particularly distressing (*P5, male: “What I fear most is… one day my personality changing… If that happened, the feeling that ‘I’m no longer myself’ would be very distressing.”)*.

Anticipated consequences of dementia were often framed in both personal and social terms, including loss of independence, inadequate care, financial strain, and fear of institutionalization. These concerns were shaped by broader social narratives about ageing, caregiving burden, and mistrust toward formal care arrangements. Such expectations, combined with uncertainty about the future, gave rise to a wide range of emotional responses. While some participants adopted a relatively accepting stance, many reported persistent worry, anxiety, and distress triggered by everyday memory lapses. As one participant described: *“I once forgot to turn off the stove while boiling water… At that moment, I was so scared that my heart was pounding” (P2, female)*.

To sum up, these findings suggest that FoD is experienced not only as concern about cognitive decline itself but also as anxiety about its potential implications for identity, autonomy, and family relationships.

### Coping with dementia-related concerns

Participants described a range of coping strategies to manage concerns about dementia, including both proactive self-management and more avoidant or emotion-focused approaches.

Many participants engaged in active self-management strategies aimed at maintaining cognitive and physical functioning. These included deliberate cognitive exercises, physical activity, and lifestyle adjustments, which were often described as ways to regain a sense of control and reduce anxiety. For example, one participant noted: *“In the morning I try to memorize a phone number. It makes me feel that my brain is still working, and that reduces my fear a bit” (P3, female)*.

At the same time, coping also involved strategies to manage emotional distress. Distraction and keeping busy were frequently mentioned, with participants engaging in household tasks or physical activities to divert attention from intrusive thoughts about memory loss. However, some participants also described more avoidant responses, such as postponing medical consultation or deliberately setting aside dementia-related concerns when feeling overwhelmed. As one participant explained: *“When these worries first came up, my initial reaction was avoidance. I pretended nothing was wrong and posted short videos to suppress” (P5, mal)*.

Beyond individual efforts, coping was shaped by access to personal and social resources. Participants’ perceived skills, particularly in obtaining health information and using digital tools, influenced their ability to seek out coping strategies and maintain a sense of competence. In addition, support from family members and peers played a crucial role in alleviating FoD, providing reassurance and opportunities to share concerns.

Overall, these findings suggest that coping with FoD involves a dynamic interplay between efforts to maintain control, regulate emotional responses, and draw on available personal and social resources.

### Support needs and preferences

Participants highlighted a range of needs, preferences, and perceived barriers and facilitators regarding potential interventions, reflecting both informational and practical considerations for engagement.

A central and recurrent theme was the need for clear, accessible, and actionable information about dementia. Participants emphasized the importance of understanding how to distinguish normal age-related memory changes from pathological decline, as well as receiving practical guidance on lifestyle and prevention. As one participant noted: *“It would be good to introduce information about dementia. At the very least, people should have a correct understanding of the disease” (P12, female)*. Another added: *“Things like what to eat, how to sleep, whether to exercise… as long as it’s explained in a way I can understand, that’s enough” (P2, female)*. Interventions were perceived as particularly valuable when they provided concrete, actionable strategies rather than abstract or fear-based messages (*P3, female: “Don’t just talk about how scary dementia is. You need to tell me what I can actually do to prevent it.”)*.

Engagement with interventions was also shaped by a dynamic balance between motivation and perceived capability. When interventions were seen as relevant, feasible, and supportive, participants expressed a strong willingness to participate, often linked to their desire to regain a sense of certainty, control, and cognitive autonomy. However, this willingness was contingent on their perceived ability to engage with the intervention, particularly in the context of digital delivery. While some participants felt confident in using smartphones and accessing online information, others reported that unfamiliar technologies or complex procedures could discourage participation (*P11, male: “When it comes to high-tech things, if the operation is complicated and hard to learn, that could indeed be a problem.”)*.

In addition, participation was influenced by broader contextual and social factors. Practical constraints such as limited time, reduced physical stamina, sensory limitations, and literacy barriers were frequently mentioned. For example, one participant noted: *“Older adults’ eyesight isn’t good… I can’t see small text at all. Older people need very large fonts” (P14, female)*. Social influences also played a key role, with participants emphasizing the importance of emotional and practical support from family members, as well as opportunities for peer interaction in non-stigmatizing environments. Trust emerged as another critical factor, with participants expressing a preference for professionally endorsed interventions and credible information sources (*P11, male: “Guidance from professionals is definitely helpful. When we try to understand things online by ourselves, the information isn’t always comprehensive or accurate. Self-diagnosing is really just amateur.”)*. Some also noted that small incentives could further encourage participation among older adults *(P15, female: “Actually, some older adults are more willing to participate if there are small gifts or incentives. That does make a difference for some, some people.”)*

Overall, participants preferred support that was credible, actionable, emotionally reassuring, and adaptable to their functional abilities and everyday circumstances. Digital delivery was acceptable to many participants but was viewed as one possible mode of support whose usefulness depended on usability, trust, and digital confidence.

## Discussion

### Principal Findings

This study extends qualitative understanding of FoD by examining not only what people fear about dementia, but also how they respond to these concerns and what forms of support they perceive as meaningful. Three findings were particularly salient. First, FoD extended beyond concern about memory loss to anticipated threats to autonomy, identity, dignity, and reciprocal family relationships. Second, participants described both proactive and avoidant coping responses, suggesting that fear may motivate efforts to maintain cognitive health while also contributing to disengagement or delayed help-seeking. Third, support needs centered on reducing uncertainty, maintaining a sense of control, and accessing credible, practical, and socially supported forms of guidance. Together, these findings position FoD as a future-oriented and relational experience whose consequences depend partly on how individuals interpret and respond to perceived cognitive threat.

### FoD as a Threat to the Future Self and Social Relationships

Our findings both converge with and extend previous qualitative work on dementia-related anxiety. Maxfield and colleagues found that community-dwelling adults without dementia described anxiety in relation to anticipated loss of self, independence, control, and future reliance on others (Maxfield, Peckham, & James, 2023; Maxfield, Peckham, James, et al., 2023). Similar concerns were evident in the present study, suggesting that dementia-related fear may be rooted in threats to the anticipated future self rather than memory loss alone. However, participants in our study also situated these concerns within family relationships, particularly the possibility of becoming dependent on adult children or disrupting established patterns of reciprocity (Dai et al., 2023).

The relational dimension was particularly salient. Participants often described future dependency in terms of its consequences for adult children and established patterns of reciprocity within the family. FoD was therefore not solely an individual appraisal of future illness but was intertwined with anticipated changes in social identity, family roles, and the ability to contribute to close relationships. These findings extend previous accounts of dementia-related anxiety by illustrating how threats to the future self may also be understood through anticipated changes in relational roles.

These relational concerns should be interpreted within, rather than attributed exclusively to, the sociocultural context of ageing in China. Expectations surrounding intergenerational care and filial support coexist with many older adults’ desire to remain independent and avoid imposing emotional, practical, or financial burdens on their children (Ng et al., 2016; Nie, 2015). Our findings therefore do not suggest that concerns about dependency or family burden are uniquely Chinese; similar concerns have been reported elsewhere (Dai et al., 2023). Rather, they indicate that the meaning of dependency may be shaped by how autonomy, reciprocity, and family responsibility are negotiated within particular social contexts.

### FoD as Both a Motivator and Barrier to Action

A key extension of previous work is the finding that FoD was associated with markedly different coping responses. Some participants attempted to preserve a sense of agency through cognitive activity, physical exercise, lifestyle adjustment, information seeking, or planning for future care. Others responded through distraction, postponement, emotional suppression, or avoidance of professional consultation. These contrasting responses suggest that fear does not lead uniformly to preventive action.

One possible explanation lies in perceived control(Kapeller & de Boer, 2024). When participants believed that meaningful action was available, concern could prompt self-management and efforts to maintain cognitive health. When dementia was perceived as uncontrollable or when confronting the possibility of dementia became emotionally overwhelming, avoidance could provide short-term relief. FoD may therefore operate as both a motivator and a barrier to action, depending on how individuals appraise the threat and their capacity to respond to it.

This distinction has practical importance. Encouraging greater awareness of dementia risk does not necessarily lead to adaptive health behavior and may inadvertently intensify anxiety or avoidance if people are left uncertain about what they can realistically do. Communication about cognitive ageing and dementia prevention should therefore balance risk information with feasible actions while avoiding messages that imply that individuals can fully control whether dementia develops.

### Support Needs: From Information to Manageable Uncertainty

Participants’ support preferences further suggest that FoD should not be approached solely as a knowledge deficit. Although participants consistently wanted reliable information, they valued information primarily when it helped them distinguish normal cognitive ageing from concerning change, reduced uncertainty, and provided feasible actions. In this sense, informational support may be most useful when it also strengthens a realistic sense of control. Emotional reassurance and relational support were also important. Family members and peers could help normalize concerns and reduce distress, while professional guidance was valued for credibility and clarity. These findings point toward support approaches that combine accurate information with emotional, behavioral, and relational components.

Moreover, older adults’ physical health status and functional capacities, such as visual limitations or reduced mobility, may further influence how individuals engage with potential interventions, and which formats they prefer. Interventions that overlook these bodily and functional needs risk excluding individuals with complex health conditions from meaningful participation (Ghosh et al., 2025). As discussed earlier, social support may also play a supportive role in facilitating engagement, suggesting that interventions addressing FoD could benefit from acknowledging the broader social context in which individuals interpret and manage their concerns.

Digital delivery may offer one feasible route for providing support, particularly where access to specialist services is limited. However, participants’ accounts suggest that digital delivery should be treated as a mode of access rather than the intervention itself. (Dai, Aardoom, et al., 2025; Dai et al., 2026; Katz et al., 2024; Moll Van Charante et al., 2024). Usability, readability, trust, digital confidence, and access to human guidance were important conditions for engagement (Badr et al., 2024; Dai, Aardoom, et al., 2025; Ghosh et al., 2025; Knox et al., 2026; Lyles et al., 2021). Offering both digital and non-digital options may therefore be more appropriate than assuming a single delivery format will suit all users.

### Methodological Reflections

The use of a third-person vignette also warrants methodological reflection. Beginning with the fictional experience of “Auntie Li” provided participants with an indirect entry point into a potentially sensitive and stigmatized topic before inviting them to relate these concerns to their own lives. This approach may have facilitated discussion by creating psychological distance and reducing the immediate pressure of personal disclosure. However, the study was not designed to evaluate the vignette as a data collection technique, and its effects on disclosure cannot be established from these data. Future qualitative gerontological research could examine whether vignette-based approaches are particularly useful for exploring emotionally difficult or stigmatized concerns.

### Implications for Practice and Intervention Development

For practice, the findings suggest that dementia-related concerns should be acknowledged rather than dismissed as an inevitable part of ageing or addressed solely through risk education. Community and ageing services could provide clear guidance on expected cognitive ageing, indicators that warrant professional consultation, and feasible strategies for supporting cognitive health. Such guidance should strengthen agency without overstating individuals’ control over dementia risk.

Future interventions may benefit from combining credible information with emotional reassurance, practical strategies, and opportunities for relational or professional support. Flexibility is also important: participants differed in physical capacity, literacy, digital confidence, coping preferences, and willingness to discuss dementia-related concerns. Intervention development should therefore remain responsive to these differences rather than assuming a uniform pathway from fear to preventive action.

### Strengths and Limitations

This study provides an in-depth account of FoD among middle-aged and older adults in China and extends previous qualitative work by examining experiences, coping responses, and support needs within the same analytical framework. The combination of Framework Analysis and the TDF supported systematic comparison across participants while retaining data-driven subthemes, and the third-person vignette offered a potentially useful approach to discussing a sensitive topic. Several limitations should be considered. The sample was relatively small and predominantly urban, which may limit the transferability of findings to rural or socioeconomically disadvantaged populations. Participants had varying levels of FoD rather than clinically defined anxiety, and the findings should therefore not be interpreted as representing individuals with severe dementia-related anxiety. Finally, although the vignette may have facilitated discussion, the study was not designed to determine whether it increased disclosure or reduced stigma.

### Conclusions

FoD among middle-aged and older adults in China was experienced as more than concern about future memory decline. It reflected anticipated threats to autonomy, identity, dignity, and family relationships and was managed through both proactive and avoidant coping responses. These findings extend existing qualitative work on dementia-related anxiety by showing how fear is negotiated in everyday life and by identifying support needs related to uncertainty, realistic control, reassurance, and accessible guidance. Understanding these processes may inform more responsive approaches to supporting dementia-related concerns in ageing populations.

## Data Availability

All data produced in the present study are available upon reasonable request to the authors

