## appendix 1 for "Fear of Dementia among Middle-Aged and Older Adults in China: Lived Experiences, Coping, and Support Needs"

### Supplementary Material 1. Semi-Structured Interview Topic Guide and Third-Person Vignette

The semi-structured topic guide was developed around the three research questions and used flexibly during data collection rather than as a fixed interview script. The English version below presents the main questions and prompts from the original Chinese topic guide used during the interviews. Open-ended questions were followed by neutral probes according to participants’ responses. A brief third-person vignette was used as an initial conversational prompt before participants were invited to discuss their own views and experiences.

#### Opening

- Could you briefly tell me about yourself and your everyday life?
- When you hear about dementia or Alzheimer’s disease, what comes to mind?

#### Third-person vignette

Auntie Li has recently noticed occasional changes in her memory. She sometimes wonders whether these changes are simply part of getting older or could mean something more serious. This makes her somewhat concerned, but she is not sure how to understand or respond to these changes.

- What are your thoughts about Auntie Li’s situation?
- What do you think she might be thinking or feeling?
- What do you think she might do?

*Participants were subsequently invited to move from the fictional scenario to their own views and experiences, if they felt comfortable doing so.*

#### Topic 1. Experiences and perceptions of fear of dementia

- Have you ever had concerns about developing dementia or about changes in your memory? Could you tell me about them?
- What does the possibility of developing dementia mean to you personally?
- What tends to make these concerns stronger or weaker?

**Possible neutral probes:** Could you tell me more about that? Can you give me an example? How did you feel at that time?

#### Topic 2. Coping with dementia-related concerns

- When concerns about memory or dementia arise, how do you usually respond?
- What, if anything, helps you manage these concerns?
- Are there times when you prefer not to think or talk about them?
- Do you ever seek information or support from other people? If so, could you tell me about that?

#### Topic 3. Support needs and preferences

- What kinds of information or support, if any, would be helpful for people who experience these concerns?
- What would make such support useful or meaningful to you?
- What might make it easier or more difficult for you to use or participate in such support?
- How would you prefer such support to be provided?

#### Closing

- Is there anything else about dementia-related concerns or support that you think is important but that we have not discussed?
