## appendix 3 for "Fear of Dementia among Middle-Aged and Older Adults in China: Lived Experiences, Coping, and Support Needs"

**Supplementary Material 2. Illustration of the Deductive–Inductive Analytical Process**

The examples below provide a simplified illustration of how selected participants’ accounts were considered during the hybrid deductive–inductive Framework Analysis. Lower-level analytic codes were developed through close engagement with participants’ accounts and considered alongside the a priori TDF domains. Codes, domain assignments, and subthemes were iteratively compared and refined within and across cases during development of the analytical framework. The examples are illustrative rather than exhaustive and do not represent a strictly linear coding sequence.

| **Research area** | **Illustrative data excerpt** | **Inductive analytic code** | **TDF-informed higher-level category** | **Final subtheme** | **Cross-case interpretation** |
| --- | --- | --- | --- | --- | --- |
| RQ1: Experiences and meanings of FoD | *“What I fear most is not losing to others, but one day my personality changing… If that happened, the feeling that ‘I’m no longer myself’ would be very distressing.” (P5)* | Anticipated loss or change of self | Social/professional role and identity | Threats to personal identity and dignity | FoD involved anticipated disruption to continuity of self and personhood, extending beyond concern about memory performance. |
| RQ1: Experiences and meanings of FoD | *“After my mother became ill, I later learned that this condition can be hereditary. Seeing her like that made me start to worry a little.” (P6)* | Personal exposure to dementia; perceived familial risk | Knowledge | Experiential and informational exposure to dementia | Direct experience and acquired knowledge shaped how participants interpreted their own future susceptibility. |
| RQ2: Coping with dementia-related concerns | *“In the morning, I try to memorize a phone number. It makes me feel that my brain is still working, and that reduces my fear a bit.” (P3)* | Deliberate cognitive exercise to reassure oneself | Behavioral regulation | Active self-management strategies | Proactive behaviors could function not only as perceived prevention but also as a way of maintaining agency and reducing fear. |
| RQ2: Coping with dementia-related concerns | *“When these worries first came up, my initial reaction was avoidance. I pretended nothing was wrong…” (P5)* | Avoiding or suppressing dementia-related concerns | Behavioral regulation | Avoidant/passive coping strategies | Avoidance could provide temporary emotional distance when dementia-related concerns felt difficult to confront. |
| RQ3: Support needs and preferences | *“Don’t just talk about how scary dementia is. You need to tell me what I can actually do to prevent it or improve things—that’s what would motivate me to participate.” (P3)* | Seeking concrete actions to reduce or manage risk | Goals | Ability to take concrete preventive actions | Participants valued support that translated dementia-related concern into feasible actions, thereby strengthening a sense of agency and control. |
| RQ3: Support needs and preferences | *“If distance is inconvenient, online options are fine too… neither format can fully replace the other.” (P4)* | Preference for flexible delivery according to circumstances | Environmental context and resources | Accessibility and availability | Acceptable delivery depended on participants’ circumstances rather than a universal preference for either digital or in-person support. |

**Abbreviation:** TDF = Theoretical Domains Framework.
